# Evaluation of 3-D printed swabs for respiratory sampling and testing for SARS-CoV-2 during the early pandemic period

**DOI:** 10.1101/2023.06.14.23291367

**Authors:** Zahra Hasan, Angila Iqbal, Imran Ahmed, Moiz Khan, Kauser Jabeen, Nazneen Islam, Erum Khan, Saleem Sayani

## Abstract

Appropriate collection of respiratory samples is essential for accurate diagnostic testing of respiratory pathogens such as, SARS-CoV-2. Early in the pandemic, there was a shortage of nasopharyngeal (NP) swabs and difficulty in sampling suspected cases. Therefore, we developed a 3D printed nasal swab for anterior nares, paired with in-house viral transport medium (VTM). The utility of this 3D swab kit was investigated in comparison with the standard NP commercial swab and VTM, in 200 individuals between August and September 2021. Subjects were those presenting for diagnostic testing for SARS-CoV-2 using the RT-PCR (cobas Roche assay) assay. NP samples were taken from each subject using the standard NP and 3D swabs followed by RT-PCR on paired specimens. CT values for amplification of gene targets were evaluated to determine assay parameters based on viral load cut offs of ≤ CT 35 or, ≤ CT 37. For high to medium viral loads, 3D swab based PCR testing had a sensitivity of 93%, specificity of 99%, positive predictive value (PPV) of 98.5% and negative predictive value (NPV) of 96.2% respectively. For low viral loads, 3D swab testing had a sensitivity of 88%, specificity of 99%, with a PPV of 98.5% and NPV of 93.2%.%. 3D swab sampling of anterior nares was comparable with NP sampling using standard swabs for SARS-CoV-2 specimens with a medium to high viral load. Therefore, 3D swab based sampling is a reliable and convenient local solution for collecting respiratory samples for SARS-CoV-2 diagnostic testing.

## Background

The global outbreak of a novel coronavirus causing severe acute respiratory distress syndrome 2019 (COVID-19) was declared a Public Health Emergency of International Concern on 30 January 2020 (1) and a pandemic on March 11, 2020, by the World Health Organization (WHO). According to John Hopkins Coronavirus Resource Center https://github.com/CSSEGISandData/COVID-19), as of 10 March 2023, SARS-CoV-2 related infections have surpassed over 765 million cases, with 6.71 million deaths globally. To date (1 April 2023), Pakistan has reported an estimated 1.57 million COVID-19 cases from 30.6 million SARS-CoV-2 PCR tests (https://ourworldindata.org/coronavirus-testing). Five successive COVID-19 waves occurred between 2020 and 2022; March and July 2020, October 2020 and January 2021, April and May 2021, July and September 2021 and, between December 2021 and February 2022 (2, 3). We focus here on the period of the first pandemic wave, which was caused mainly by the Wuhan and G clade lineage strains (4).

Rapid diagnosis of infectious organisms such as respiratory viruses is dependent on accurate detection of the pathogen in an appropriate sample. Routine testing is done typically by a nasopharyngeal swab is inserted in the nostril to collect secretions from the upper respiratory tract to diagnose respiratory infections such as, SARS-CoV-2. For respiratory viruses such as, influenza and SARS-CoV-2 which initiate infection in the upper respiratory tract, collection of samples from the nasopharynx, oropharynx and nasal cavities have all been shown to be effective for virus diagnostics. Samples are collected and placed in a viral transport medium (VTM) that prevents virus degradation so that nucleic acid testing can be conducted. Nasopharyngeal (NP) swabs are usually flocked flexible swabs made of alignate, or polyester, that are routinely imported into Pakistan. NP swabs were in limited supply in particularly during the first six months of the pandemic (March until August 2020) when lockdowns were in place, making it difficult to import items into Pakistan.

The unavailability of NP swabs was a big bottleneck for the collection of respiratory specimens for SARS-CoV-2 testing. Further, appropriate transport medium for collection and storage prior to downstream PCR was required. To address this bottleneck for SARS-CoV-2 testing, there was a worldwide effort by university academic medical centers and commercial companies to initiate local manufacture of swabs through 3D printing devices. Formlabs was one of the first institutes to meet the global high demand of nasal swabs for testing of COVID-19. University of South Florida (USF) health faculty and Northwell Health validated the implementation of 3D printing nasal swabs. Results from their testing have demonstrated that the 3D printed nasal swabs perform equally well in comparison to standard swabs, for COVID-19 nasopharyngeal testing (5, 6).

In Pakistan, the Aga Khan University Hospital (AKUH), Karachi, Pakistan was the first private sector laboratory to introduce SARS-CoV-2 RT-PCR testing in February 2020. AKUH was at the front-line of COVID-19 diagnostics during the pandemic but soon faced a shortage of respiratory virus sampling kits. Therefore, AKU developed 3D printed swab with an in-house viral transport medium (VTM) for respiratory sampling. Further, respiratory sample collection was conducted using anterior nares (nasal) sampling for additional convenience. Here we describe the clinical validation of 3-D printed swabs for collection of nasal samples in a laboratory developed universal transport medium for SARS-CoV-2 PCR testing. The study compared the results of swabs testing using both NP and 3D swab methods followed by RT-PCR testing of each for SARS-CoV-2 RNA.

## Methods

### Ethical and Regulatory considerations

This study received approval from the Ethical Review Committee, Aga Khan University, Pakistan. It was also approved by the Clinical Trials Unit, AKU and conducted in accordance with relevant regulations of Good Clinical Practice.

### Subject Selection and consent

Study subjects recruited were those coming for routine diagnostic testing to the AKUH laboratory COVID-19 testing zone from August 5– September 5, 2020. Male or female adults aged 18 – 65 years were included in the study. Exclusion criteria were those who did not submit both NP and 3D swab samples for testing. Study participants were offered an incentive of Pakistani rupee 500, to be reduced from their routine COVID-19 PCR test bill for participation in this study.

### Printing of 3D swabs detail

The Formlabs’ Form 3a Printer uses Surgical Guide Resin material to print the bristle prototype nasal swabs. It is a CE certified, biocompatible Photopolymer Resin material, which meets Class I requirements (5–7). The 3D nasal swab made at the Aga Khan University is designed leveraging learnings from USF and other globally available software designs. The swab was 3D printed on Formlab’s Form 3a SLA printer and washed using isopropyl alcohol (IPA) in Formlab’s wash (8). Each swab is dried and finely tuned to enhance its mechanical properties using Form Cure. Post-curing, swabs can be chemically disinfected or steam sterilized in an autoclave for final packaging (9), Figure 1.

**Figure 1.**
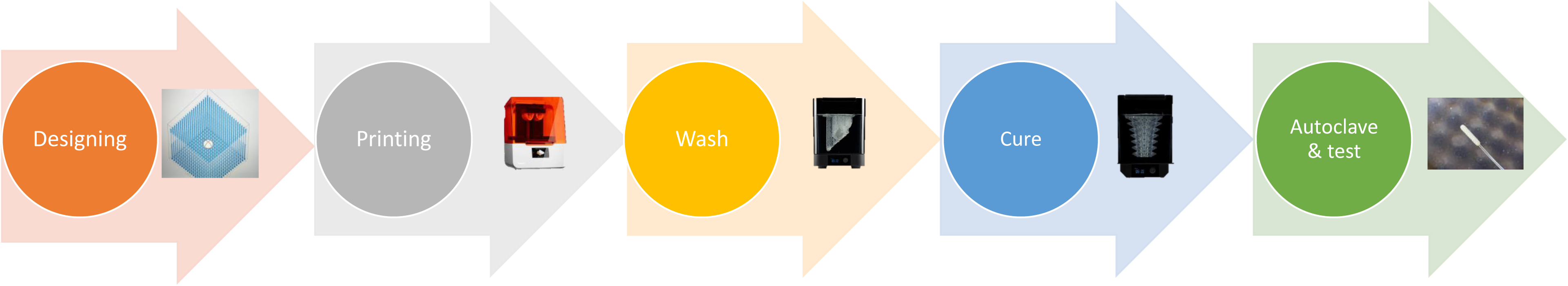
Process of Nasal Swab Printing

The swabs underwent a comprehensive evaluation process, with five distinct design variations denoted as D1 to D5. These designs incorporated minor structural modifications. D1, D2 and D3 were nasopharyngeal (NP) type 3D swabs. D4 and D5 were nasal type 3D swabs. Clinical testing was carried out on each design to evaluate their performance. The NP type of 3D swabs (D1-D3) were found to be uncomfortable by the study subjects. Therefore, we focused on D4 and D5. D4 was found to be more fragile whilst D5 was robust and effective for collection of nasal swabs for PCR testing.

A clinical trial involving a sample size of 200, was conducted with the most promising design, D5. A patent application (#US17/127,604) was filed in the USA (10) for the 3D printed nasal swab in December 2020.

### Preparation of universal transport medium

Laboratory developed VTM was prepared for the collection and storage of respiratory samples using guidelines available from the World Health Organization (WHO). Media was prepared as per standard protocol for a Phosphate-Buffered-Saline-glycerol medium containing antibiotics. The PBS-glycerol VTM was autoclaved and then poured into sterile receiver tubes in an aseptic manner in the Clinical Microbiology media room. The media was also checked for sterility and its ability to inhibit growth of bacteria. The media was subsequently stored at 4°C for 1–2 days or, at -20°C for longer term storage. Validation of the VTM was conducted as per CAP guidelines (Supplementary Files 1-2).

### Sampling kits

The 3D printed swab was used to collect specimens in the in house VTM (Figure 2). The commercial swab used was a nasopharyngeal FLOQ (NP) swab with VTM from Beaver, USA.

**Figure 2.**
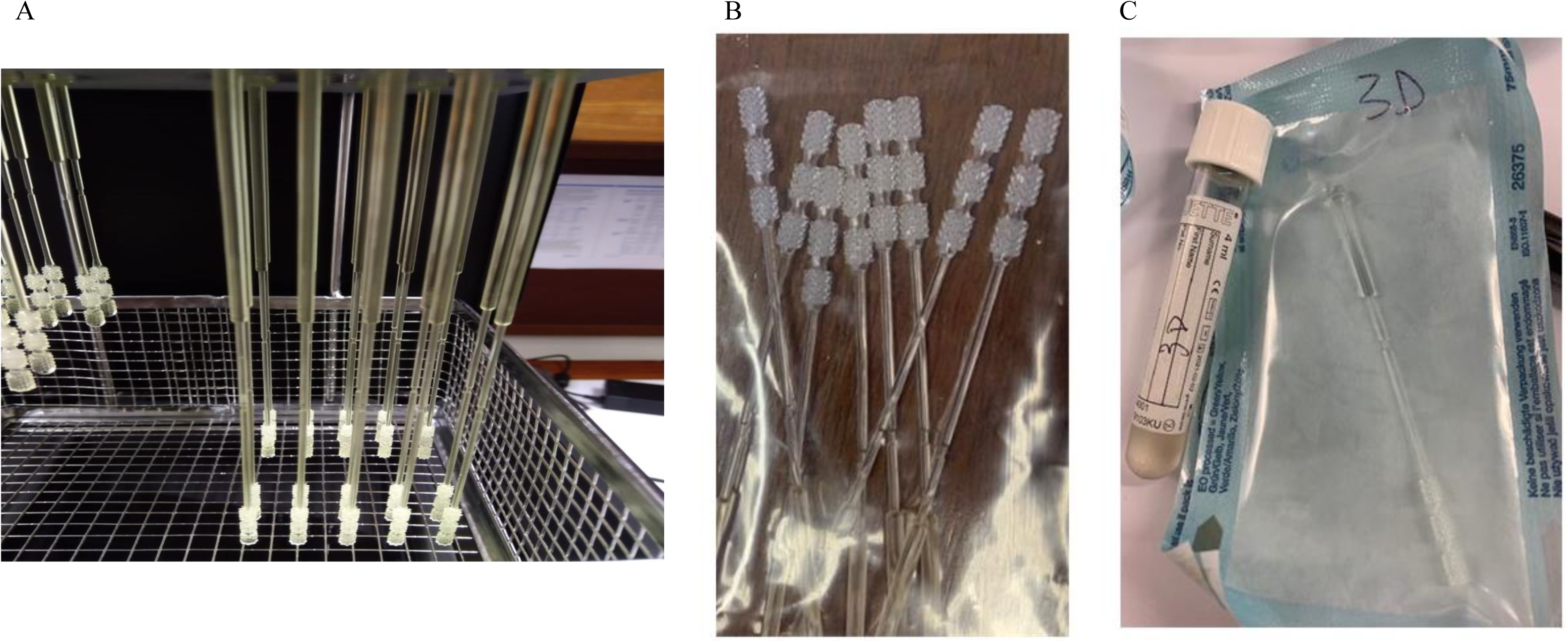
3D nasal swab detail. A, View of the 3D swab during the drying process. B, Packaged 3D swab. C, Autoclaved nasal swab with viral transport medium.

### Sample size calculation

At the time the study was initiated in August 2020, the SARS-CoV-2 PCR positivity prevalence rate was 20-25%. For accuracy testing according to College of American Pathologists (CAP), USA guidelines we needed to have at least 20 positive samples. For this purpose, we set the sample size to recruit 200 individuals for the study.

### Study design

This was a validation study comparing NP and 3D printed swab based respiratory sampling method used for SARS-CoV-2 RT-PCR testing. It involved consecutive convenience sampling of individuals for up to 10 individuals per day. A research officer was responsible for recruitment of study subjects, taking consent and for ensuring transportation of the study samples to the laboratory for PCR testing.

Each individual was given an instruction sheet to guide collection of the nasal sample. After the guidance, they were asked to collect their sample using the 3D swab kit and submit it for SARS-CoV-2 PCR testing. The self-sampling was observed by the study research officer. The study participants were also asked to give their feedback on their experience of using the 3D swab to self-collect samples. Responses were documented on the data form.

### RT-PCR testing

Respiratory swab specimens were tested for SARS-CoV-2 using the Cobas^®^ SARS-CoV-2 RT-PCR assay (Roche Diagnostics, USA). Real time RT-PCR for SARS-CoV-2 Orf1 and Envelope (E) gene targets.

Based on results of individuals PCR for each specimen, a positive or negative result was determined in each case using cut offs CT > 35 or, CT <= 35 respectively. For each specimen, this was interpreted based on analysis that allowed Chi Squared analysis based on whether samples were true positive (TP), false positive (FP), true negative (TN) or false negative (FN).

### Analytical plan

Demographic data including age and gender, and clinical and laboratory information was documented. Data on study participants and their test results was kept confidential and anonymous. For each individual, the results of the SARS-CoV-2 PCR assay based on NP swab sampling would be compared with the results obtained using the 3D nasal swab sampling method. Data were used to determine sensitivity, specificity, positive and negative predictive value of the assay. Further, questions related to ease of sampling were asked of the study participants. These were compiled into qualitative results.

## Results

### Pilot study comparing sampling with 3D nasal swab/VTM with standard NP/VTM kit

We first conducted a pilot study in volunteer healthcare workers, recruiting 19 laboratory staff and healthcare workers who consented to participate in sampling with the NP swab followed by the 3D nasal swab. The individuals were first sampled using the NP method and then performed a self-collection using the 3D nasal swab as per instructions. These subjects comprised 16 healthy individuals with no signs and symptoms of COVID-19. Three were those who had tested positive for COVID-19 in the prior 24–48 h (this was done in a quarantine setting). The healthy individuals without symptoms were considered negative controls and those who had a positive SARS-CoV-2 PCR test were considered positive controls. The 16 healthy individuals all tested negative for SARS-CoV-2 after 3D swab sampled PCR testing (Supplementary Table 1). The three individuals with a prior positive COVID-19 test were positive after 3D swab-based sampling and PCR testing. Therefore, there were 16 true negative and 3 true positive test results, resulting in a test sensitivity and specificity of 100 percent for the 3D nasal swab/VTM based sampling.

### Comparison of 3D nasal swab and standard NP swab collection for SARS-CoV-2 testing in a prospective cohort

We conducted a trial of diagnostic testing of individuals who presented to the institutional COVID-19 testing zone for SARS-CoV-2 RT-PCR testing. We recruited 218 individuals to the study, comprising 129 males and 72 females. The average age of study participants was 41.2 years. Sixteen individuals were excluded from the final analysis due to either test cancellation or failure to submit a comparative sample. Data for 202 individuals was available for comparative analysis of 3D swab VTM swab and commercial NP VTM swab sampling for RT-PCR (Figure 3). In each case, the result of the PCR test was compared both qualitatively and quantitatively. Therefore, PCR test results of both NP and 3D nasal swabs were compared using the parameters of the laboratory validation.

**Figure 3.**
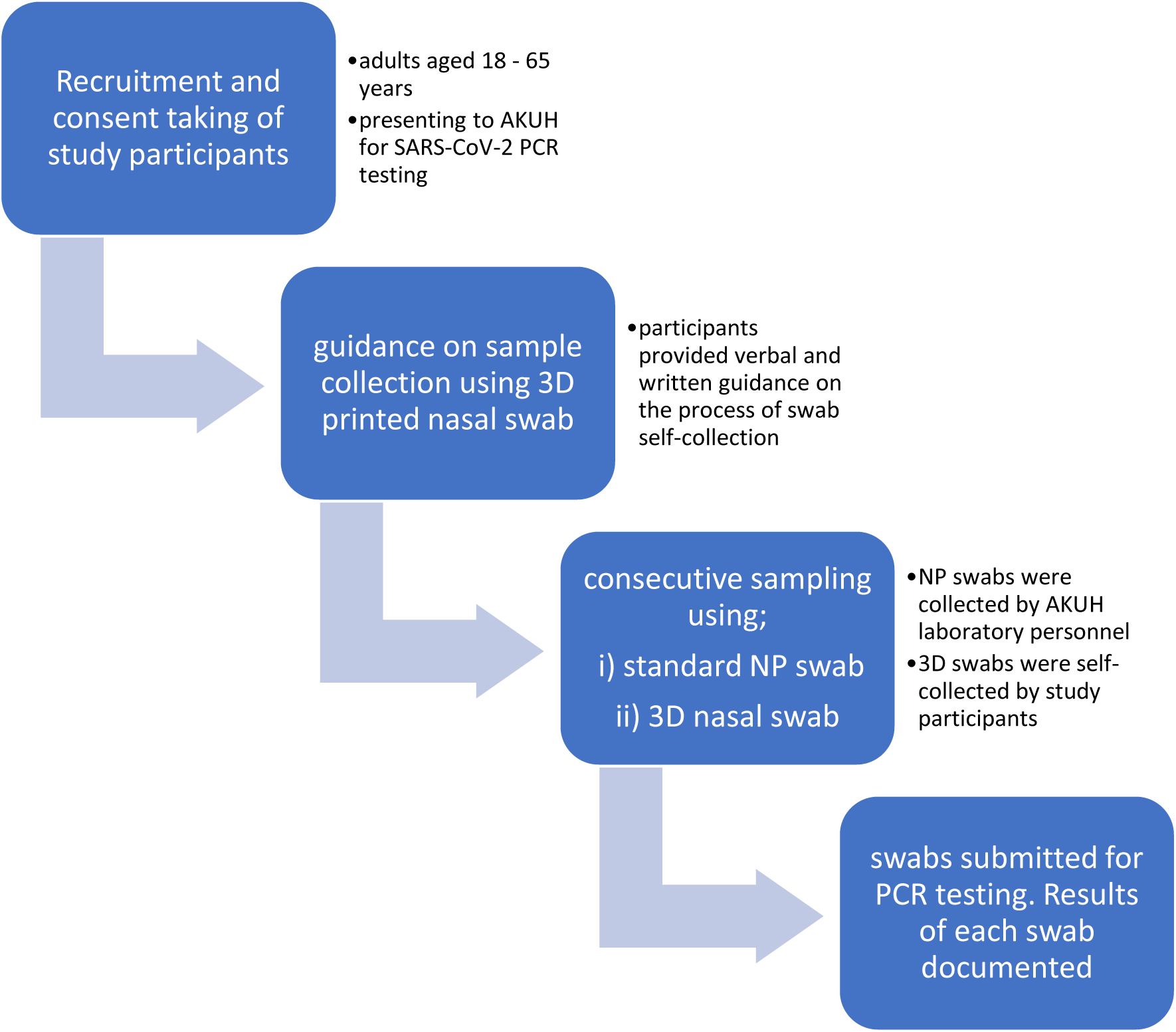
Study plan for comparing 3D printed nasal as compared with commercial nasopharyngeal swab for SARS-CoV-2 PCR testing

Comparative PCR results of the NP and 3D swab methods are shown in Figure 4. There was no difference in the diagnostic CT value of Orf1 gene amplification for the 200 specimens testing using either NP or 3D swabs. Similarly, CT values for E gene amplification in the 202 specimens tested with either NP or 3D swabs was equivalent. A Spearman’s rank correlation analysis was run between the diagnostic PCR between both swabs for the two gene targets. There was a strong correlation between both methods for Orf1 gene detection (rho 0.6738, with p value 1.43 e^9^). E gene target PCR using both methods was also strongly correlated (rho 0.638, p 8.01 e^9^).

**Figure 4.**
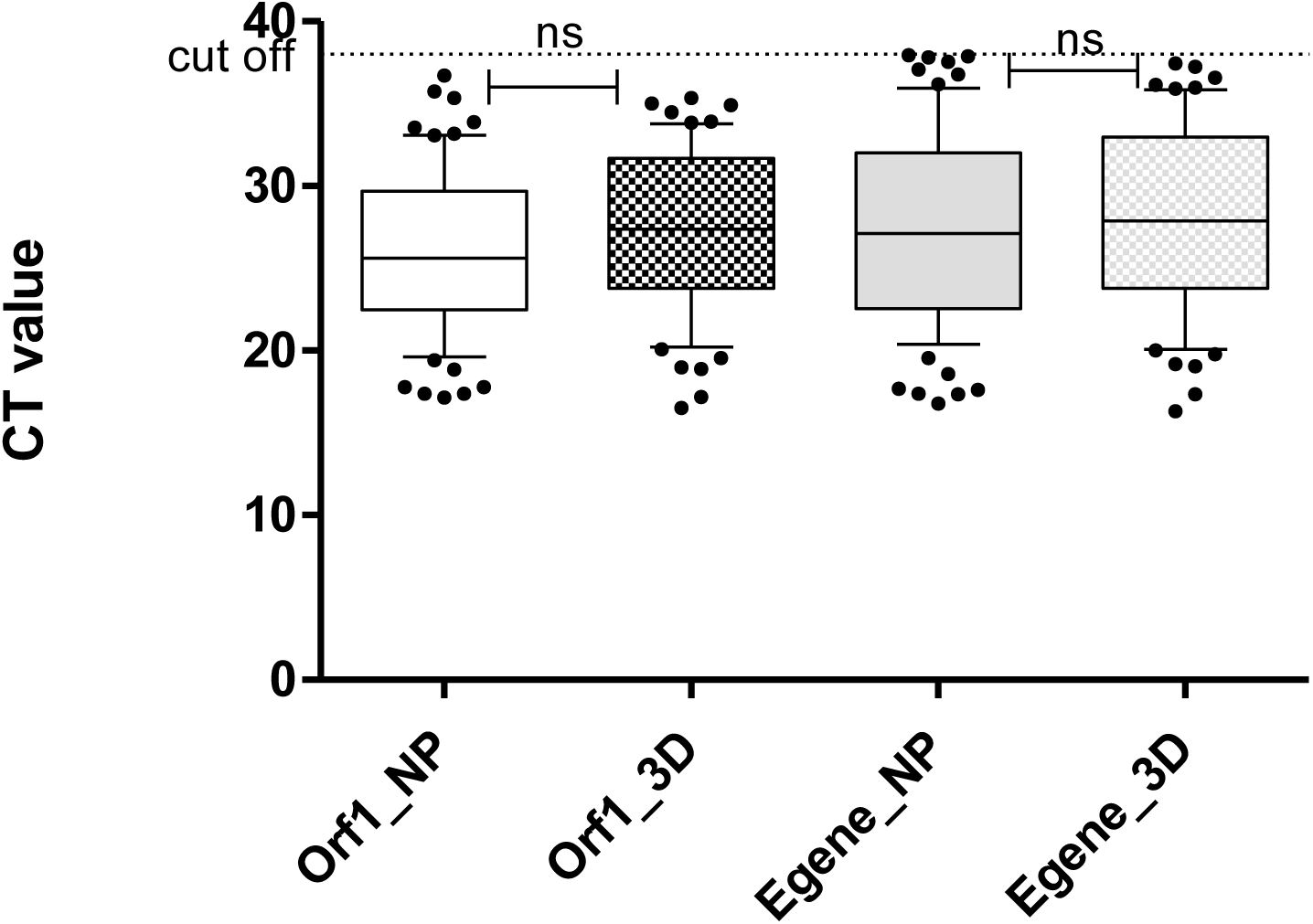
PCR based testing of specimens using NP and 3D swabs was comparable. Graphs depict the SARS-CoV-2 RT-PCR result in CT values for the Orf1 and Envelope (E (gene targets in the SARS-CoV-2 RT-PCR cobas assay. Results of Positive samples detected based on Orf1 and E gene detection are compared between NP and 3D swab specimens. The CT values of each PCR result is illustrated. Statistical comparison between CT values for each gene target between the swab types was conducted using the Mann Whitney U assay, ns denotes p>0.05, ns, not significant.

### Assessment of sampling experience using the 3D nasal swab

Qualitative data on using the swab was available for 107 study participants. Of these, 87.9% found it easy to use whilst 10.2% found it difficult (Table 1). The remaining two individuals did not collect the samples themselves and preferred that the laboratory staff to conduct it.

**Table 1.** Assessment of ease of self-sampling with a 3D printed swab

|  | Total data<br>available<br>(N) | Frequency<br>of total<br>group (%) |
| --- | --- | --- |
|  | 107 |  |
| Very easy # | 94 | 87.9 |
| Difficult/did<br>not like it | 11 | 10.2 |
| Other<br>comments * | 2 | 1.9 |

Responses regarding ease of performing self-sampling using a 3D swab are provided. \*sampling was performed by laboratory staff at request of participants. # included data from 17 healthcare workers

### Sensitivity and Specificity testing of the 3D swab kit method

Of the 202 individuals tested using the NP swab kit, 75 were Positive and 127 were Negative for SARS-CoV-2 viral RNA (Supplementary Table 2). Testing using the NP swab kit was used as the Gold standard method. Results of the 3D swab kit-based PCR were compared against the gold standard in two parts. The first was using the SARS-CoV-2 RT-PCR diagnostic assay validated cut-off of CT 37. This was the limit of detection (LOD) established by the AKUH laboratories as per CAP guidelines. A CT 37 would identify samples of a low viral load. In this manner, 3D swab based testing identified 66 positive specimens in the total group.

The second option was to use a cut-off of CT 35 for identification of a positive SARS-CoV-2 RT-PCR test. A CT of 35 would capture samples of a high to medium viral load. Here, 3D swab based testing identified 72 positive specimens.

We determined the sensitivity and specificity of our 3D swab based PCR testing in the context of both cut offs, at CT 37 and CT 35. Figure 5 depicts the results for comparison of 3D and NP swab sampling for specimens based on both the cut offs. When a cut off CT 37 was used, PCR based testing using 3D swabs had a sensitivity, 88% and specificity 99%. This resulted in a positive predictive value (PPV) of 98.5% and a negative predictive value (NPV) of 93.2%.

**Figure 5.**
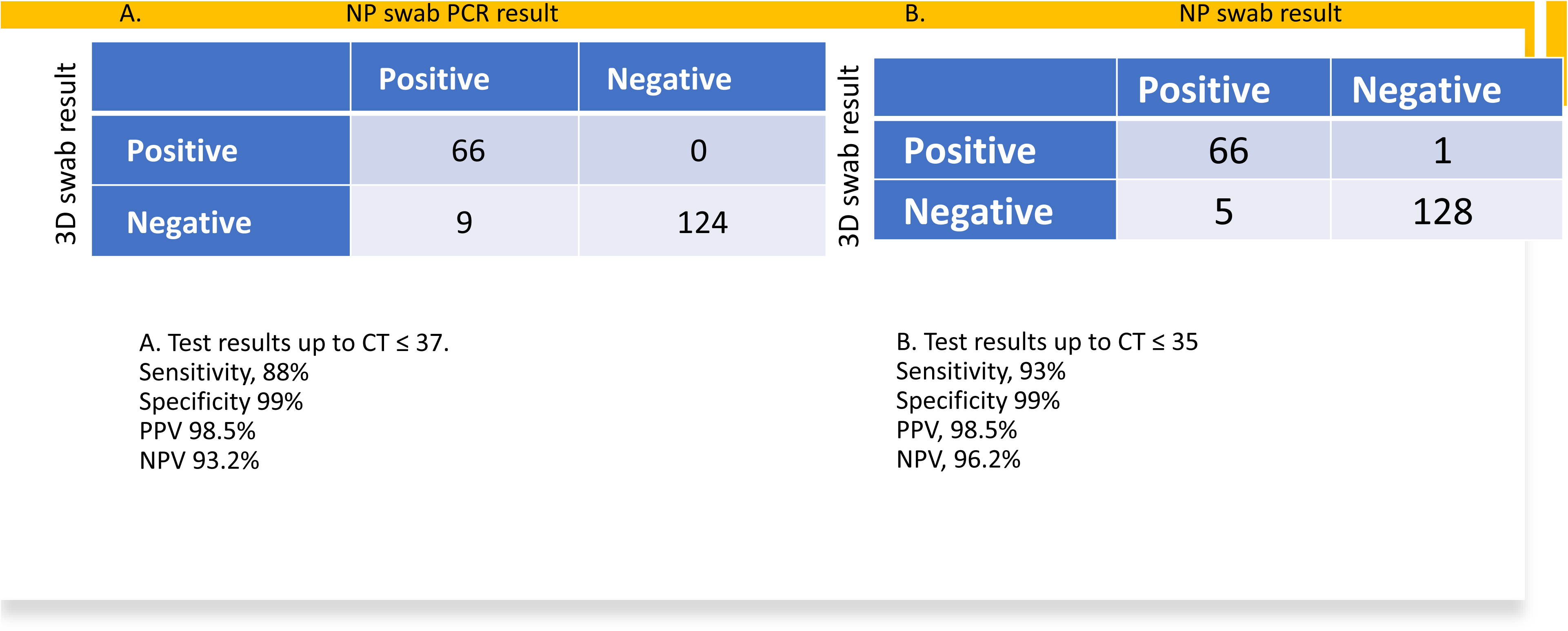
Sensitivity and Specificity calculation of 3D and NP swab PCR results. The tables below interpret PCR results of specimens tested from each participant using either the NP or the 3D swab. Results are interpreted based on True Positive, False Positive, True Negative or False Negative results used to determine sensitivity, specificity, PPV and NPV for each assay. Cut offs for CT used were that or the Orf1 gene in each case at either A, CT ≤ 37 or B, CT ≤ 35.

When a cut off 35 CT was used, 3D swab based PCR testing had a sensitivity of 93%; specificity of 99%; PPV of 98.5% and NPV of 96.2%.

## DISCUSSION

Our validation study data demonstrates robust sensitivity and specificity of an in-house manufactured 3D swab in comparison with the standard NP commercial swab for amplification of the Orfl and E gene targets. In using nasal swabs instead of nasopharygeal swabs, it also broke the bottleneck related to complex sampling for collection of specimens, allowing self-collection of specimens. The comparative results of the 3D swab results with the NP swab revealed high sensitivity and specificity particularly, for high to medium viral load specimens. With, a slightly reduced sensitivity in the case of low viral load load specimens.

3D printing of nasal swabs provide a novel solution for overcoming a global dilemma to meet the increasing demands for COVID-19 testing through rapid design reproducibility coupled with manufacturing at high throughput levels. This was successfully demonstrated in April 2020 by Callahan et al at Beth Israel Deaconess Medical Center, who with the collaboration of health care workers and 3D printing companies manufactured and validated four prototypes of 3D nasal swabs in a span of only 22 days (12).

Previous studies exploring the effectiveness of in-house 3D printed swabs showed similar results (13–16). A study from Canada in 2021 validating a 3D swab manufactured with Surgical Guide Resin in samples from sixty three participants using the Cobas Roche assay found a high concordance with the standard NP swab with positive predictive value (PPV) of 96.8% and negative predictive value (NPV) of 96.9% (13). Furthermore, in a study conducted at two national hospitals of Singapore in 2021, Tay et al. reported a positive percentage agreement (PPA) of more than 91% between a 3D printed swab and the reference swabs (FLOQ and polyester Dacron swabs) using the Cobas Roche assay for amplification of Orflab and E gene targets in samples from seventy nine patients (14).

In our cohort of patients, 3D nasal swabs showed a lower sensitivity when a cut off > 35 CT was used as compared to when a cut off ≤ 35 CT was used. The CT value generated based on PCR amplification of gene targets in SARS-CoV-2 has been shown to be inversely correlated with the viral load of clinical specimens. Hence, low viral load samples are those with high CT values, and vice versa. Further, data understanding the transmissibility of SARS-CoV-2 specimens and also their viability has shown that viral load specimens up to CT 35 are those with a likelihood of transmission (11). It has been demonstrated by previous studies that the concordance between nasal and NP swabs depends on Limit of detection (LoD) of the PCR assay with higher concordance for higher viral loads as compared to low viral loads. As lower viral loads may lie below the LoD therefore, may result in false negative results. A study from Boston, USA conducted in June 2020 compared CT values from RT-PCR testing in nasal and NP swabs collected at the same time from three hundred and eight clinically suspected COVID-19 cases. A high concordance among the nasal and NP samples was found only with viral loads above 1000 copies/ml whereas, lower concordance was seen in samples with viral loads below 1000 copies/ml, most of which were missed when nasal swabs alone (17).

The 3D nasal swabs at our center were manufactured with Surgical Guide Resin material modeled on the bristle prototype. Several studies evaluating the material of different 3D printed swabs and their compatibility with the RT-PCR testing workflow as well as concordance of SARS-CoV-2 testing results with the standard NP FLOQ swabs, have demonstrated favorable results regarding this prototype (18, 19). Furthermore, the 3D-printed swabs can be adjusted for flexibility, shaft length and diameter during the manufacturing process thereby allowing for easy adaptation as compared to FLOQ swabs. Although the Formlabs 3D-printed swabs we used for our study are less flexible as compared to elastic materials of other 3D printers, the Formlabs 3D printers are cost effective and easily employed in hospitals with their own 3D printing laboratories (18).

Self collection of samples is a breakthrough for COVID-19 testing as it provides a convenient, safe and compliant approach. A large number of the participants (87.9%) found it easy to use the 3D printed swab in our study. In agreement with studies from other centers, this approach has been found to be non-inferior to the observed collection of nasal swab specimens and hence, should be encouraged for better compliance for disease detection and screening (20, 21).

Our study had a few limitations. Firstly, this was a single center study and validation data from other centers from Pakistan are required for establishing the use 3D printed swabs all over Pakistan. Nonetheless, Aga Khan University is one of the largest tertiary care hospitals in Pakistan and receives patients from areas representative from all over the country. We focused on the 3D swab prototype which was a nasal type. We did not use other 3D swab materials in our study. However, the study participants found our bristle 3D swab prototype easy and convenient to use and has provided a stepping stone for the evaluation of other swab types and materials in future.

To the best of our knowledge this is the first study from Pakistan validating an in-house printed 3D swab for COVID-19 testing and comparing it with the standard NP swab. These results highlight the significance of ongoing refinement and innovation in medical device design to optimize clinical performance and enhance patient care. It also demonstrates how new initiatives can be tested in a local setting, removing dependence on external supplies.

## Data Availability

All data produced in the present work are contained in the manuscript

## Acknowledgments

We thank the Aga Khan University Hospital Clinical Laboratories and Department of Pathology and Laboratory Medicine, teams of phlebotomists and Technologists for their contributions.

